# The Development, Optimization, and Validation of Four Different Machine Learning Algorithms to Identify Ventilator Dyssynchrony

**DOI:** 10.1101/2023.11.28.23299134

**Authors:** Peter D Sottile, Bradford Smith, Marc Moss, David J Albers

## Abstract

**Objective:** Invasive mechanical ventilation can worsen lung injury. Ventilator dyssynchrony (VD) may propagate ventilator-induced lung injury (VILI) and is challenging to detect and systematically monitor because each patient takes approximately 25,000 breaths a day yet some types of VD are rare, accounting for less than 1% of all breaths. Therefore, we sought to develop and validate accurate machine learning (ML) algorithms to detect multiple types of VD by leveraging esophageal pressure waveform data to quantify patient effort with airway pressure, flow, and volume data generated during mechanical ventilation, building a computational pipeline to facilitate the study of VD.

*Materials and Methods:* We collected ventilator waveform and esophageal pressure data from 30 patients admitted to the ICU. Esophageal pressure allows the measurement of transpulmonary pressure and patient effort. Waveform data were cleaned, features considered essential to VD detection were calculated, and a set of 10,000 breaths were manually labeled. Four ML algorithms were trained to classify each type of VD: logistic regression, support vector classification, random forest, and XGBoost.

*Results:* We trained ML models to detect different families and seven types of VD with high sensitivity (>90% and >80%, respectively). Three types of VD remained difficult for ML to classify because of their rarity and lack of sample size. XGBoost classified breaths with increased specificity compared to other ML algorithms.

*Discussion:* We developed ML models to detect multiple types of VD accurately. The ability to accurately detect multiple VD types addresses one of the significant limitations in understanding the role of VD in affecting patient outcomes.

*Conclusion:* ML models identify multiple types of VD by utilizing esophageal pressure data and airway pressure, flow, and volume waveforms. The development of such computational pipelines will facilitate the identification of VD in a scalable fashion, allowing for the systematic study of VD and its impact on patient outcomes.

## BACKGROUND AND SIGNIFICANCE

Invasive mechanical ventilation is a life-saving intervention. But mechanical ventilation can damage the lung, termed ventilator-induced lung injury (VILI).^1^ VILI is caused by large tidal volumes, high pressures, or repeated alveolar collapse and worsens patient outcomes. Low tidal volume ventilation (LTVV) is one method to reduce VILI and improve outcomes.^2–6^ Yet, the complex interaction between the patient and the ventilator limits the efficacy of a one-size-fits-all LTVV strategy. Defined as the inappropriate timing and delivery of a breath in response to patient effort, ventilator dyssynchrony (VD) limts the effect of LTVV and potentiates VILI.^7^

However, VD and its impact are difficult to monitor and quantify for many reasons. First, the average patient receiving mechanical ventilation receives over 25,000 breaths daily. Historically, nearly 40% of ICU admissions require mechanical ventilation, resulting in millions of mechanically ventilated breaths daily in a single hospital - far too many for manual evaluation and necessitating the need for automated detection.^8^

Second, multiple types of VD can impact the patient in several ways. The risk of high-volume or high-pressure ventilation varies by the type and severity of VD.^9–12^ Significant variation within a given type of VD may represent differences in severity and the potential to propagate VILI. Moreover, even these prototypical definitions of VD present with substantial breath-to-breath variation, further complicating identification (Figure 1).

**Figure 1:**
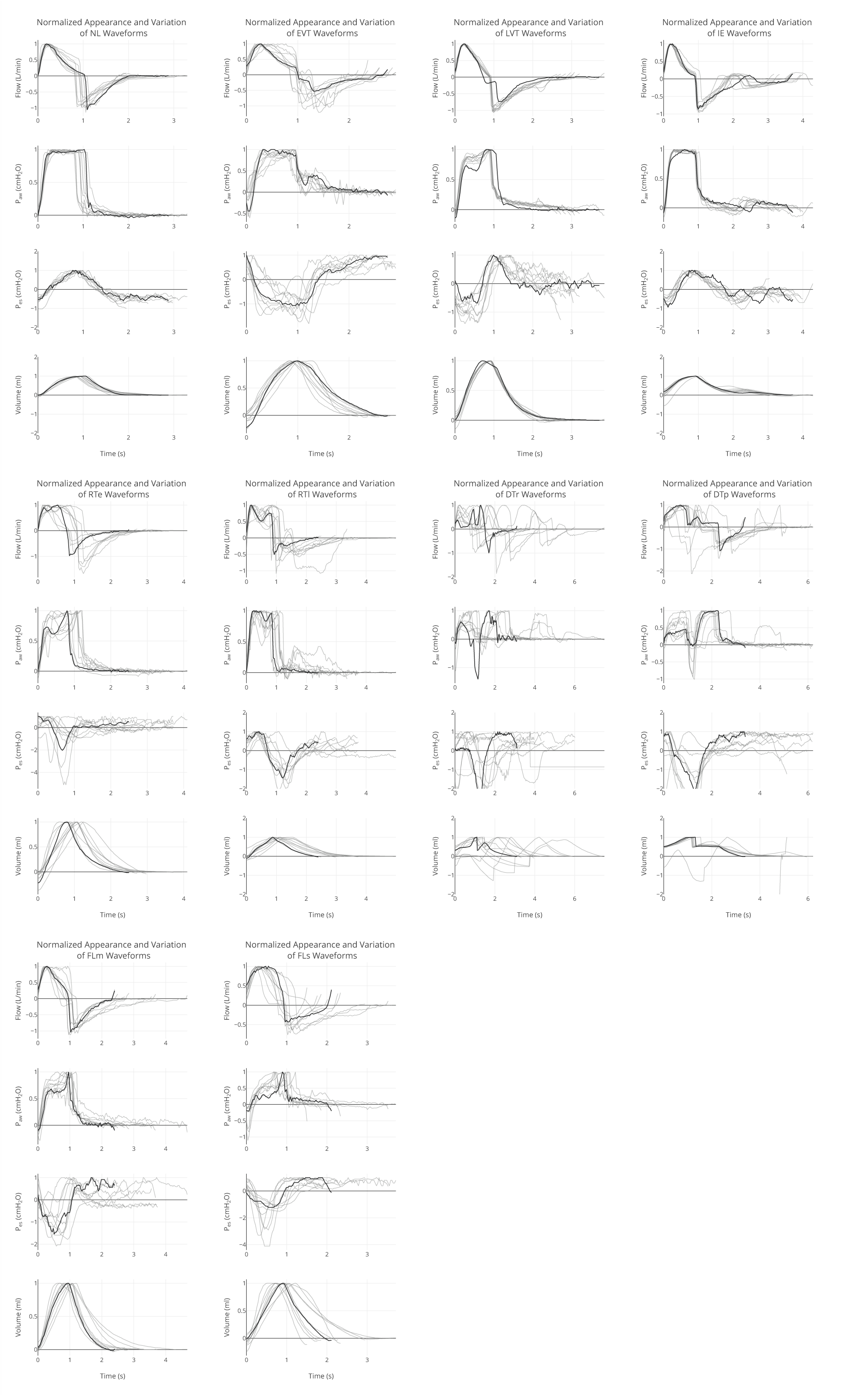
Examples Images of Types of Ventilator Dyssynchrony. Prototypical example of each type of VD and 10 examples of the variation observed in each type of VD; **NL**: normal breath, **IE:** ineffective trigger - characterized by a negative deflection of the Pes during expiration; **FLm**: mild flow-limited - a negative Pes deflection during inspiration with coving of the Paw waveform; **FLs**: severe flow-limited - a negative Pes deflection during inspiration with more pronounced coving of the Paw waveform; **LVT**: late ventilator termination - the inspiratory cycle is terminated during active expiration; **EVT**: early ventilator termination - the inspiratory cycle is terminated before active inspiration is complete; **DTp**: patient-triggered double-triggered - a second breath is initiated by patient effort before complete expiration; **RTe**: early reverse-triggered - a reflexive contraction of the diaphragm that starts after passive insufflation of the lung but before the end of inspiration, **RTl**: late-reverse triggered - a reflexive contraction of the diaphragm that starts after a passive insufflation of the lung but after the inspiratory cycle is complete; **DTr**: reverse-triggered double-triggered - a double-triggered breath that is starts with a passive insufflation of the lung triggered a reflexive diaphragmatic contraction that continues into expiration and triggers a second breath before expiration is complete.

Third, several types of dyssynchrony can only be accurately detected with additional monitoring, such as esophageal manometry.^10,13,14^ While we and others have previously described automated methods to detect a subset of VD utilizing standard airway pressure and flow waveforms, the detection of additional types of VD that require advanced monitoring remains difficult.^9,15–23^ These advanced monitoring techniques, such as esophageal manometry, require expertise for placement and have nuanced interpretation.^24^ Integrating these advanced monitoring techniques into automated VD detection algorithms is necessary to detect all types of VD in critically ill patients.

Finally, VD’s frequency and severity are heterogeneous between patients and over time in a given patient.^25–28^ Some types of VD are rare, accounting for less than 1% of the breaths a patient takes and may be clustered in time, therefore generating an irregular and sparse dataset that complicates classification. Correllating the potential of VD to worsen VILI is further complicated by change in the patient’s underlying condition over time. Thus, it is necessary to study VD throughout a patient’s entire course to understand its effects on VILI - analysis of a subset of breaths is not sufficient to understand these interactions.

## OBJECTIVE

The inability to label VD breaths automatically at scale impairs our ability to reduce VILI due to VD and little work has been done in this scope until recently. Furthermore, delineating the association between VD and VILI requires the development of an accurate VD detection algorithm that does not necessitate manual monitoring, can scale to multiple patients and millions of breaths, and can integrate with the EHR to allow extraction of patient-specific data that varies over time.

We sought to develop a computational pipeline to identify multiple VD types accurately, integrate esophageal manometry, and compare the results using four supervised machine learning (ML) techniques. Multiple ML techniques were evaluated because it is difficult to predict which ML technique will perform best for a given data set, particularly when the outcomes of interest are rare.^29^ This study represents a computational pipeline to facilitate future studies to reduce ventilation-related morbidity and mortality.

## MATERIALS AND METHODS

### Data Collection

We included patients admitted to the University of Colorado Medical, Surgical, Neurosurgical, and COVID ICUs. Patients between 18 and 89 years old, needing mechanical ventilation, and with Acute Respiratory Distress Syndrome (ARDS) by the Berlin definition or ARDS risk factors were enrolled within 24 hours of intubation.^30^ Alternatively, patients could be enrolled within 24 hours of the placement of an esophageal balloon by the clinical team. At-risk patients were defined as intubated patients with a mechanism of lung injury known to cause ARDS who have not met chest x-ray or oxygenation criteria for ARDS. All patients were ventilated with a Hamilton G5 ventilator. The Colorado Multiple Institutional Review Board (COMIRB) gave eithical approval for this study.

Patients were excluded if: a) less than 18 years of age, b) pregnant, c) imprisoned, d) with esophageal injury, recent esophageal or gastric surgery (3 months), e) tracheal-esophageal fistula, f) facial fracture, or g) active or recent (3 months) variceal bleed or banding.

Patient characteristics were extracted from the EHR. Continuous ventilator data were collected using a laptop connected to the ventilator using Hamilton DataLogger software (Hamilton, v5.0, 2011) to obtain time-stamped measures of airway pressure (p_aw_), flow, volume, and esophageal pressure measurements (p_es_). Data were recorded at 32ms intervals. Additionally, the DataLogger software collects ventilator mode and ventilator settings. Data were collected at enrollment for up to 48 hours after esophageal balloon placement to reduce the effects of length of time bias. An esophageal balloon pressure monitor (CooperSurgical; Truball, CT) was inserted into the sedated and intubated patient at the time of enrollment for all patients. The balloon occlusion test confirmed placement.

Esophageal balloon pressure monitoring allows for the direct estimation of transpulmonary pressure, which defines the pressures across the pulmonary parenchyma, independent of extrathoracic forces, and thus better defines the potential for lung injury and allows for the allow for the quantification of patient effort.^23,24^ The timing and magnitude of patient effort, or the lack thereof, helps to define the presence and severity of specific types of VD.^18,24^

### Types of VD

Eleven types of breaths were identified for this study (Table 2, Figure 1).^9,10,26^ Some of these types of VD have similar characteristics and were grouped into families of VD to simplify identification.

First, the family of normal-appearing (NL) breaths is characterized by an appearance of the airway pressure, flow, and esophageal pressure waveforms of a patient breathing on the ventilator without respiratory muscle contraction deforming the expected airway pressure and flow waveforms. Normal-appearing breaths can be passive (ventilator driven) or spontaneous (initiated by the patient).

Second, the family of reverse-triggered (RT) breaths is characterized by an initial passive breath triggering a reflexive diaphragmatic contraction during the passive breath’s inspiratory or expiratory phase.^10,31,32^ The family of reverse-triggered breaths has several different phenotypic appearances depending on the timing and strength of the reflexive contraction of the diaphragm. In early-reverse triggered (RTe) breaths, the peak reflexive contraction occurs during inspiration. In late-reverse triggered (RTl) breaths, the peak reflexive contraction occurs during expiration. Finally, in reverse-triggered double-triggered breaths (DTr), the reflexive contraction - either during inspiration or expiration - is strong enough to trigger the ventilator to give a full second breath, resulting in a double-triggered breath.^10,14^ These DTr breaths are also co-labeled in the double-triggered family of breaths. The esophageal balloon is necessary for defining the initial breath as passive (a positive deflection in the p_es_) followed by diaphragmatic contraction (a subsequent negative deflection in p_es_).

Third, the family of double-triggered (DT) breaths is characterized by a couplet of breaths. In this scenario, either patient effort or reflexive contraction of the diaphragm is strong enough to trigger the ventilator to deliver a second, full breath before the initial breath is completely exhaled. This means that two inspiratory cycles are stacked in succession without allowing time for the complete exhalation of the first inspiratory cycle. If driven by patient effort, it is a patient-triggered double-triggered breath (DTp), and a negative p_es_ deflection is seen throughout both inspiratory cycles. Auto-triggering (DTa) occurs when an artifact triggers excessive breaths.

Alternatively, the breath can be a reverse-triggered, double-triggered breath, as described above. Finally, the second inspiratory cycle in a DT couplet was identified as a post-double-triggered breath (DTpost), so that an accurately identified couplet had a DTr, DTp, or DTa cycle followed by a DTpost to complete dyssynchronus pattern.

Fourth, the family of flow-limited (FL) breaths is defined by patient effort (a negative p_es_ deflection during inspiration) exceeding the inspiratory flow achieved by the ventilator. This results in a coving of the airway pressure waveform. Breaths were empirically classified as mild (FLm) if coving was less than 50% of peak airway pressure and severe (FLs) if greater than 50% of the peak airway pressure.

Finally, the remainder of the breaths were categorized as miscellaneous and did not share common characteristics. Early ventilator-terminated (EVT) breaths are characterized by the ventilator terminating the breath before the patient ceases inspiratory effort. Late ventilator-terminated (LVT) breaths are characterized by the patient finishing inspiration before the ventilator, leading to a steep rise in airway pressure near the end of inspiration. Ineffective triggered breaths (IE) are when the patient’s effort is insufficient to trigger a breath.

### Computational Methods

A training set of 10,000 breaths from 30 patients was randomly selected for manual review and labeling by one of the investigators (PDS). The manual review of breath tracings was the gold standard to identify VD.^33^ Algorithm development followed the classic supervised-learning classification problem. The process of developing this classification model follows five steps: 1) data cleaning and verification, 2) feature engineering, 3) feature selection, 4) model for training, and 5) model validation.

#### 1. Data Cleaning

While the airway pressure, flow, and volume waveforms produced by the ventilator were interpretable in their raw form, esophageal pressure measurements required cleaning and preparation because of noise and drift in the signal. Because the location and pressure of the esophageal balloon can change with patient movement, both baseline trends and the amplitude of oscillation in the p_es_ tracing could suddenly change. Moreover, the esophageal balloon deflated slowly over time, damping the mean and amplitude. Finally, physiologic processes, namely esophageal peristalsis and cardiac oscillations, introduce cyclical signals to the baseline measurement at a frequency either much slower than each breath (i.e., 1x every 60s for esophageal spasm) or much faster than each breath (i.e., typically <1 second for cardiac oscillations).

We manually reviewed all p_es_ tracings to identify periods with adequate signal, identify time points with a sudden change in signal quality, and note periods where the balloon deflated and was then reinflated. The deflation-induced linear trend in the mean was removed for each period. Due to deflation, the best signal was at the beginning of each recording period, when the balloon was freshly inflated. Consequently, the amplitude of the p_es_ tracing for each breath during the first three minutes of a period was identified. The median of these values was then utilized to define a baseline amplitude for the entire period. The entire period was then divided into 30-second windows. If the mean amplitude of the p_es_ for a given window was decreased compared to the baseline, the window was standardized to have a mean amplitude equal to that of the baseline, preserving breath-to-breath variation related to patient effort. If the amplitude of each window was greater than the baseline, it was left unaltered, as this was thought to represent periods of increased patient effort.

To remove the high-frequency noise generated by cardiac oscillations, a low pass filter of 0.83Hz (equivalent to a heart rate of 50 beats/minute) was applied to the p_es_ waveform (Appendix, Figure A2).^34^ Esophageal spasm was relatively rare and, therefore not filtered.

Auto-triggered breaths were not included in the final ML training set, given very few examples in the manually labeled set (16 of 10,000 randomly selected breaths, Table 1). Finally, any breaths with missing data or that could not be manually labeled from the original 10,000 breaths were removed from the labeled set.

**Table 1:**
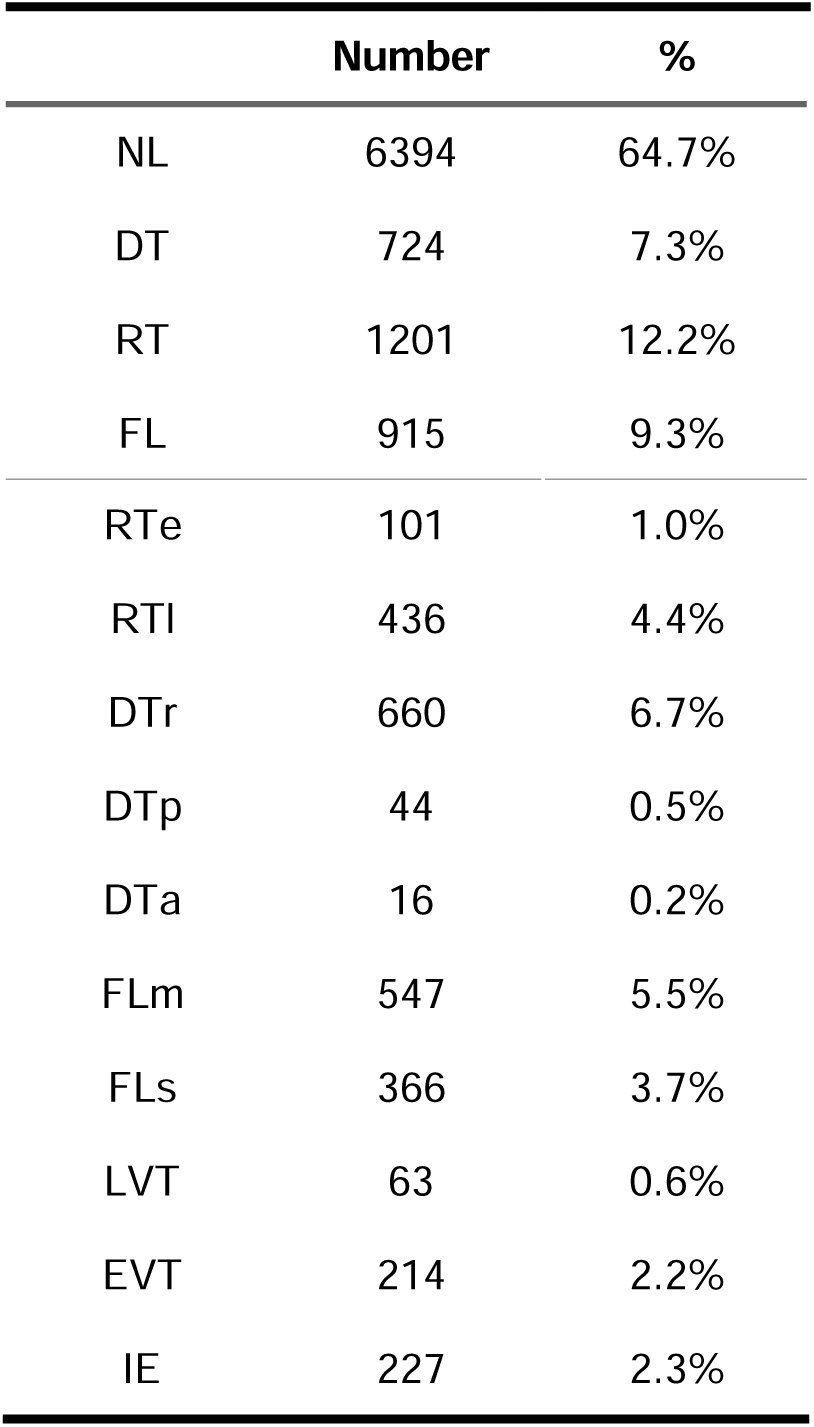
Frequency of Each Class of Dyssynchrony in Manually Labeled Set. NL: normal, **DT**: double-triggered, **RT**: reverse-triggered, **FL**: flow-limited; **RTe**: early reverse-triggered, **RTl**: late reverse-triggered, **DTr**: double-triggered reverse-triggered, **DTp**: patient-triggered double-triggered, **DTa**: auto double triggered, **FLm**: mild flow-limited, **FLs**: severe flow-limited, LVT: late ventilator termination, **EVT**: early ventilator termination, **IE**: ineffective-triggered

**Table 2:**
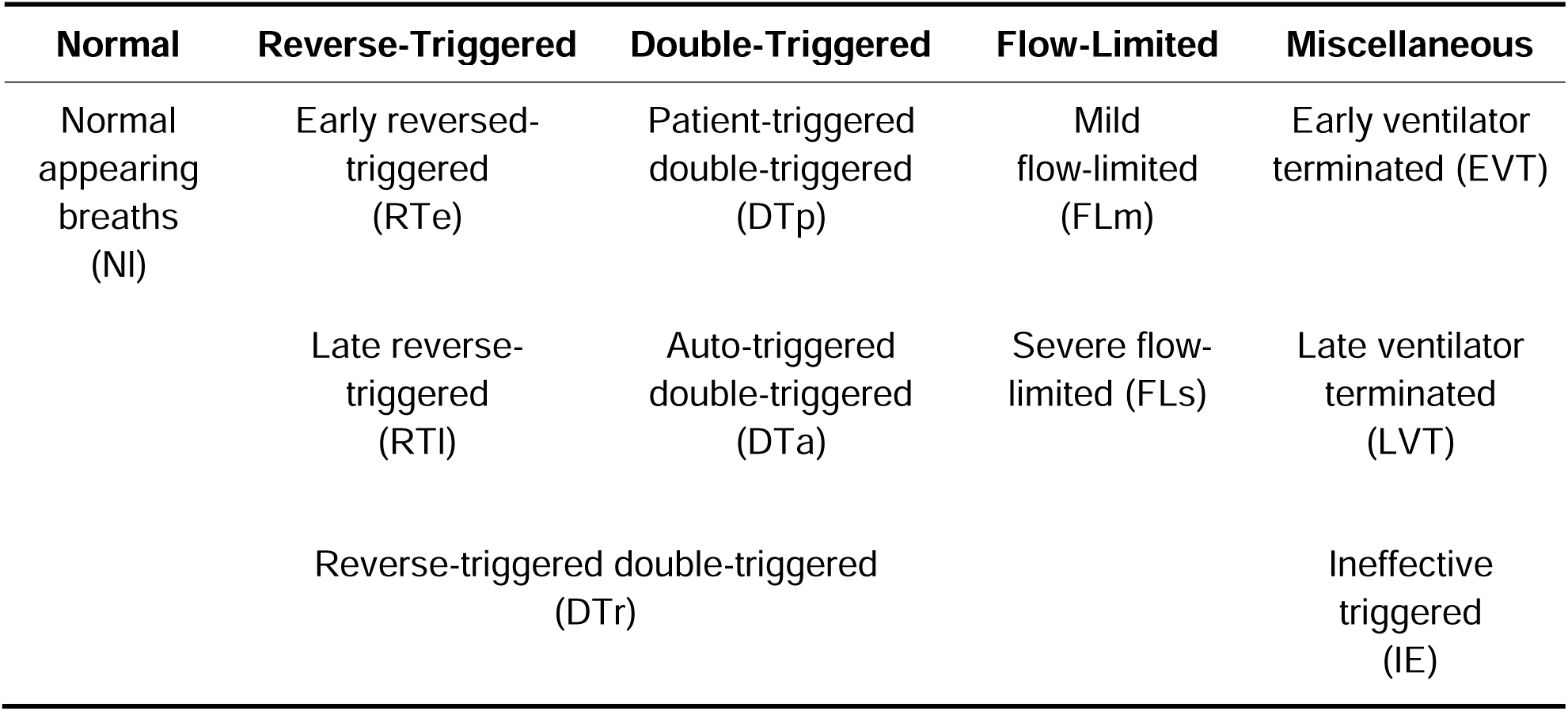
Types of VD and Their Respective Families.

| **Normal** | **Reverse-Triggered** | **Double-Triggered** | **Flow-Limited** | **Miscellaneous** |
| --- | --- | --- | --- | --- |
| Normal appearing breaths (NI) | Early reversed-triggered (RTe) | Patient-triggered double-triggered (DTp) | Mild flow-limited (FLm) | Early ventilator terminated (EVT) |
|  | Late reverse-triggered (RTl) | Auto-triggered double-triggered (DTa) | Severe flow-limited (FLs) | Late ventilator terminated (LVT) |
|  | Reverse-triggered double-triggered (DTr) |  |  | Ineffective triggered (IE) |

#### 2. Feature Engineering

We identified breath-by-breath features based on clinical expertise likely to aid in identifying VD. All airway pressure, flow, and esophageal pressure waveforms were standardized by each breath to have a maximum of one. Further, p_es_ waveforms were normalized to have a mean of zero.

Next, general features were identified. These features sought to quantify differences in waveform charactersitics between each type of VD (see Figure 1 and Figure A1). These features have been described in details elsewhere and highlight the canonical differences between each type of VD.^7,9,10,14^ These included total breath time, inspiratory time, and expiratory time, the p.10 (pressure at the first 0.1s of inspiration), and the variance in each breath’s airway and esophageal pressure.

We then identified characteristics of the two largest peaks of each inspiratory and expiratory cycle for airway pressure and flow tracings: maximum, minimum, peak location, peak width at 20% and 80% of the peak height, left intersection point at 20% and 80% of the peak height, base width, and peak prominence (Appendix, Figure A1; also see Scipy online documentation for the function scpiy.signal.find_peaks for details of these metrics).^35^

The timing of patient muscular contraction compared to inspiration and expiration is important to differentiate a reverse-triggered breath. We sought to define the lag between maximal muscular contraction (determined from the p_es_) and the peak airway pressure. We defined the lag at the maximum absolute value of the time-lagged cross-correlation between airway pressure and p_es_. The sign (positive/negative) at the time of maximum cross-correlation defined a passive or spontaneous breath, respectively. The next closest peak was selected if the maximum lag was greater than 30% of the breath length. Finally, the time between the most positive or negative p_es_ deviation and the end-inspiratory was calculated.

Then, we partitioned each breath into seven equal parts during the inspiratory phase and five equal parts during the expiratory phase. This parameterized the amplitude of the waveform at uniform times throughout each breath. The expiratory phase of a breath generally has less variability and thus needs fewer features to characterize it. The median value of each phase was calculated for airway pressure, flow, and p_es_ tracings. This effectively undersampled the breath to allow for uniform measurement of the median magnitude of each waveform (flow, airway pressure, esophageal pressure) at predefined intervals of the breath.

Finally, using the labeled training set, we calculated the average airway pressure, flow, and esophageal pressure at each measurement time for each type of VD. We then calculated the Euclidean distance of each breath’s airway pressure, flow, and esophageal pressure to the mean distance calculated from manually labeled data. This allowed us to measure how much each breath differed from the mean of each type of VD.

These calculations generated 174 individual features.

#### 3. Feature Selection

To minimize overfitting, individual features were manually pruned. Violin plots of each feature, stratified by each type of VD, were analyzed (Appendix, Figure A3). If the distribution of each feature had substantial visual overlap for each type of VD, it was eliminated from the set of possible features. Moreover, this process confirmed that the features calculated resulted in clinically logical values for each type of VD. These steps removed 39% of the features yielding a total of 107 possible features included in the final training set. Automatic feature selection was then utilized by optimizing appropriate hyperparameters to minimize overfitting.

#### 4. Model Development

Four ML algorithms were compared for VD classification: lasso logistic classification, support vector classification (SVM), random forest classification, and XGBoost classification (Figure 2).^36–38^ Importantly, types of VD are not mutually exclusive; a one-versus-all identification technique was used for identification. Moreover, because some types of VD have similar characteristics, they were first grouped into four families: normal, reverse-triggered, double-triggered, and flow-limited breaths. Consequently, a separate ML model was needed for each VD type, generating 40 independent models to evaluate four algorithms across ten VD types and three VD families.

**Figure 2:**
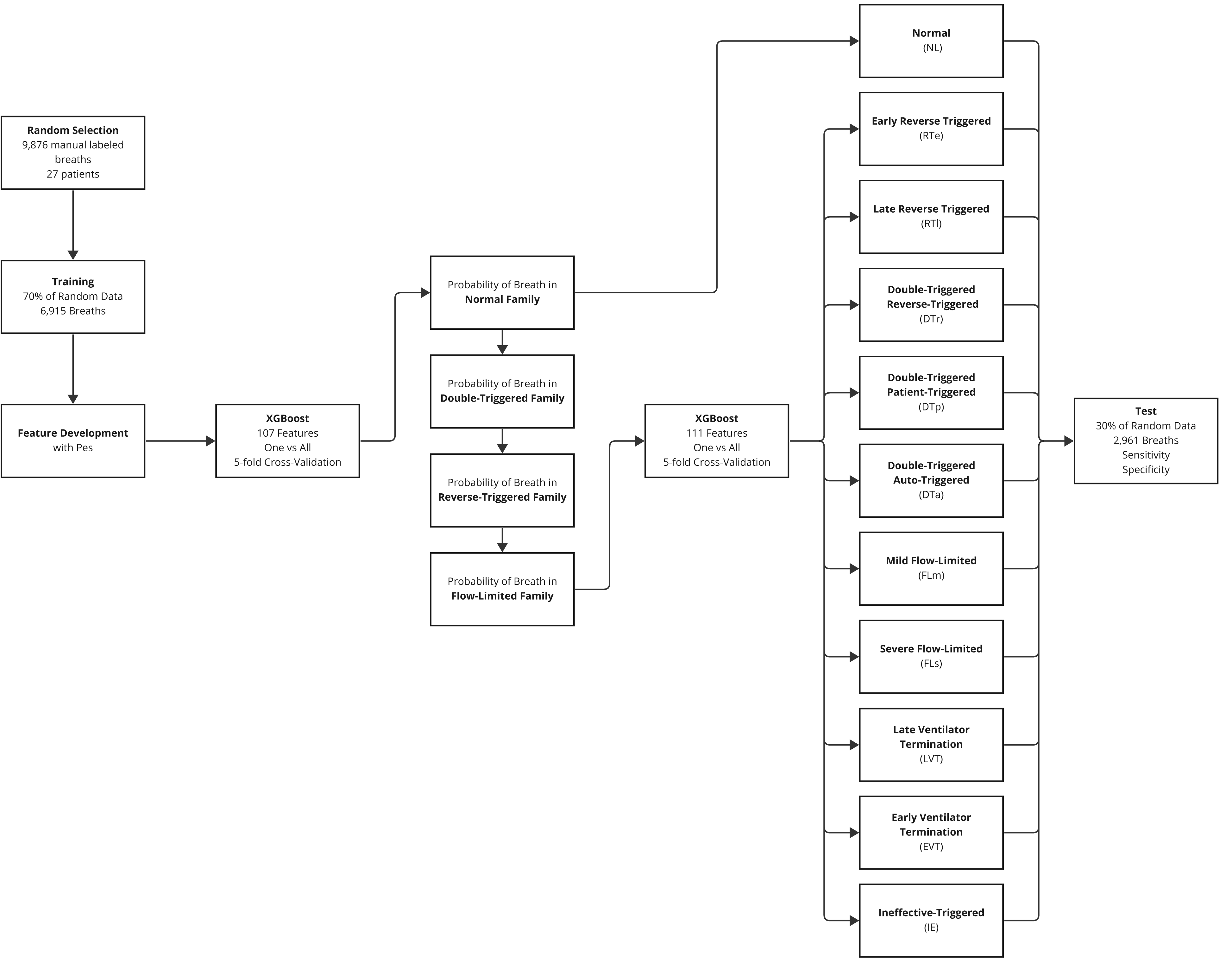
Flow Diagram of ML Training and Validation.

The same training, test, and validation strategy was used for all ML algorithms. The labeled data set was split 70:30% into training and validation sets, stratified by each class of VD. Each model was trained using 5-fold stratified cross-validation.^29^ Because categories were not evenly distributed, models were trained to maximize sensitivity. Specificity, accuracy, F1-score, false-negative, and false-positive rates were calculated as secondary outcome measures (Table 2 and Appendix, Table A1).

We sequentially estimated the probability of a breath being normal, then DT, RT, and FL. After each estimation, the probability of a given breath being in a specific family was added as a feature. Specifically, the model to identify normal breaths was trained first using the 107 features above. Next, the model to estimate the probability of a breath being double-triggered was trained with 107 original features plus the probability that the breath is normal. The probability of a breath being double triggered was added to the feature set to estimate the subsequent families - RT and FL. Consequently, when a model to predict the probability of a specific type of VD (e.g., early reverse-triggered or RTe) was trained, the feature set included the probability of the breath being normal, reverse-triggered, double-triggered, or flow-limited, in addition to the original 107 features. Importantly, the feature set did not include the probability of a breath being a specific type of VD.

Optimal hyperparameters for each ML algorithm and VD family/type were identified using nested 3-fold stratified cross-validation and a random search strategy. The lasso logistic regression hyperparameter λ was optimized between a range of 0 to 10. The SVM cost parameter *C* was optimized between 0 and 100. A radial basis function kernel was utilized for SVM classification. The parameters of the random forest classifiers max_depth and n_estimators were trained between 0 to 10 and 10 to 100, respectively. Finally, XGBoost hyperparameters were trained as follows: η 0.01 - 0.5, _ 0 -10, α 0 - 10, max_depth 4 - 10, subsample 0.3 - 1.0, min_child_weight 1-5. These parameters were optimized to reduce over-fitting. All other hyperparameters were left to their default value for all models.^29^

#### 5. Model Validation

Finally, after training, all models were validated against the 30% hold-out set, with each family and type of VD validated in the same order as the training set. Additionally, because a one-versus-all approach was utilized for VD identification resulting in 10 independent models, we analyzed trends when the models incorrectly labeled a breath. Moreover, we investigated the percentage and types of breaths that were given no labels or multiple labels.

## RESULTS

Of the original 30 patients, a total of 27 patients were included in the manually labeled training set of 9,876 breaths. Three patients had too few breaths with adequate P_es_ and were excluded. Normal breaths were the most common in the manually labeled set of breaths (64.7% of labeled breaths), while most individual types of dyssynchronous breaths were relatively rare (<10%, see Table 1.

XGBoost generally outperformed other ML algorithms in the detection of VD (Table 3). Increasing algorithm complexity (logistic classification to SVM to Random Forest to XGBoost) improved testing characteristics across all types of VD without increasing the risk of overfitting, as indicated by similar test characteristics between the CV-training and validation sets (Table 3). However, all algorithms generally identified RTe, DTp, and LVT breaths poorly and showed evidence of overfitting. Analysis of the falsely labeled breaths demonstrated different patterns for different types of VD (Figure 3). Some falsely labeled breaths appeared to be equally distributed across the types of VD (i.e., NL), while others were closely related to other types in the same family (i.e., RTe being labeled RTl). FLm breaths were mislabeled with as either RTe or LVT breaths as all three types demonstrate coving during the inspiratory phase of the airway pressure waveform.

**Figure 3:**
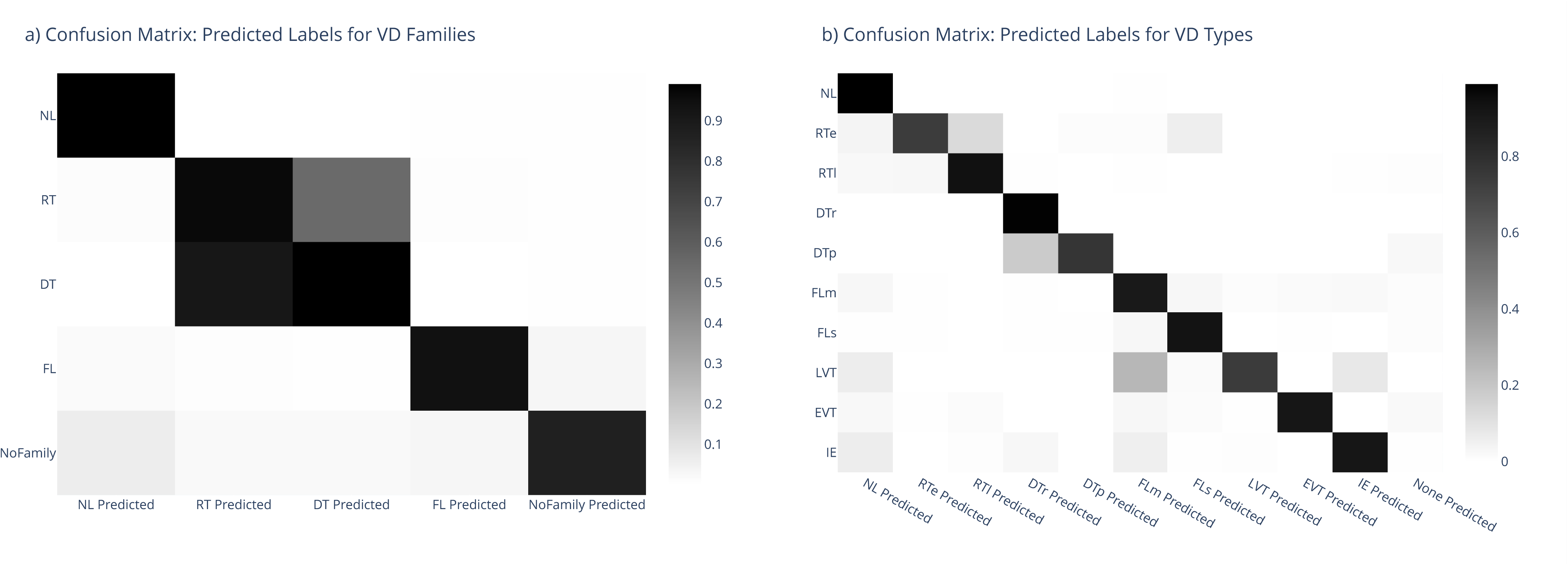
Confusion Matrix of Labeled Breaths. Percentage (grayscale bar) predicted labels correctly assigned for VD families (a) and specific VD types (b). Some RT breaths are also DT breaths, so the overlap is expected at the family level. Moreover, not all breaths have a VD family, such as (IE, EVT, LVT). **NL**: normal, **DT**: double-triggered, **RT**: reverse-triggered, **FL**: flow-limited; **RTe**: early reverse-triggered, **RTl**: late reverse-triggered, **DTr**: double-triggered reverse-triggered, **DTp**: patient-triggered double-triggered, **FLm**: mild flow-limited, **FLs**: severe flow-limited, **LVT**: late ventilator termination, **EVT**: early ventilator termination, **IE**: ineffective-triggered

**Table 3:** Test Characteristics of ML Algorithms with Esophageal Pressures: mean % ± SD; **NL**: normal, **DT**: double-triggered, **RT**: reverse-triggered, **FL**: flow-limited; **RTe**: early reverse-triggered, **RTl**: late reverse-triggered, **DTr**: double-triggered reverse-triggered, **DTp**: patient-triggered double-triggered, **FLm**: mild flow-limited, **FLs**: severe flow-limited, **LVT**: late ventilator termination, **EVT**: early ventilator termination, **IE**: ineffective-triggered

|  | Lasso Logistic Regression |  | Support Vector Classification |  | Random Forest |  |  | XGBoost |  |  |
| --- | --- | --- | --- | --- | --- | --- | --- | --- | --- | --- |
|  | Sensitivity | Specificity | Sensitivity | Specificity | Sensitivity | Validation Sensitivity | Specificity | Sensitivity | Validation Sensitivity | Specificity |
| NL | 96.6±0.6 | 80.8±4.8 | 96.7±0.4 | 93.7±1.3 | 97.2±0.5 | 97.4 | 93.6±1.6 | 97.1±0.5 | 96.3 | 95.9±0.9 |
| DT | 62.9±7.7 | 99.3±0.2 | 85.8±3.7 | 99.4±0.3 | 87.5±3.7 | 89.8 | 99.7±0.1 | 92.7±2.3 | 91.7 | 99.7±0.1 |
| RT | 54.5±14.9 | 98.7±0.4 | 86.3±2.4 | 98.9±0.3 | 88.8±3.3 | 88.2 | 99.3±0.2 | 93.4±1.4 | 90.0 | 99.2±0.2 |
| FL | 54.1±10.6 | 98.6±0.4 | 76.9±3.0 | 98.8±0.3 | 88.7±1.8 | 86.7 | 98.9±0.3 | 89.1±2.5 | 83.6 | 99.1±0.3 |
| RTe | 0.0±0.0 | 99.9±0.1 | 11.6±6.4 | 99.9±0.1 | 8.3±7.2 | 5.0 | 99.9±0.1 | 49.4±12.5 | 23.3 | 99.8±0.1 |
| RTI | 62.1±25.2 | 99.6±0.2 | 76.9±0.2 | 99.6±0.1 | 85.8±3.8 | 86.8 | 99.6±0.2 | 91.4±2.8 | 83.9 | 99.6±0.2 |
| DTr | 57.9±10.5 | 99.3±0.2 | 84.1±3.8 | 99.4±0.2 | 96.6±1.4 | 94.4 | 99.7±0.2 | 98.6±1.1 | 93.9 | 99.9±0.1 |
| DTp | 2.3±7.9 | 99.9±0.1 | 22.3±13.7 | 99.9±0.1 | 22.7±17.6 | 10.0 | 100±0.0 | 63.7±14.6 | 23.1 | 99.9±0.1 |
| FLm | 7.6±11.8 | 99.5±0.4 | 55.7±7.8 | 99.0±0.2 | 81.1±5.1 | 75.9 | 99.2±0.3 | 90.0±3.2 | 71.8 | 99.5±0.1 |
| FLs | 16.1±18.8 | 99.6±0.2 | 68.6.1±7.8 | 99.4±0.3 | 84.9±4.1 | 85.5 | 99.6±0.2 | 87.7±4.9 | 81.7 | 99.6±0.2 |
| LVT | 0.0±0.0 | 99.3±0.1 | 2.0±4.1 | 99.9±0.1 | 1.9±4.6 | 9.1 | 100±0.0 | 41.8±13.7 | 47.4 | 99.9±0.1 |
| EVT | 1.8±7.0 | 99.9±0.1 | 51.±13.3 | 99.7±0.2 | 54.9±9.6 | 58.8 | 99.9±0.1 | 70.4±6.9 | 67.2 | 99.9±0.1 |
| IE | 7.6±14.5 | 99.9±0.1 | 74.4±7.2 | 99.9±0.1 | 70.0±8.1 | 72.3 | 99.9±0.1 | 78.1±9.2 | 85.5 | 99.9±0.1 |

No XGBoost labeled breaths were inappropriately co-labeled with a mutually exclusive label. However, not all VD types are mutually exclusive - for instance IE breaths can be and were observed with other VD types. Similarly, breaths receiving no label were rare (34 breaths, 0.3%). Unlabeled breaths made up between 0 to 2.27% (DTp breaths) of each VD type (Figure 4).

**Figure 4:**
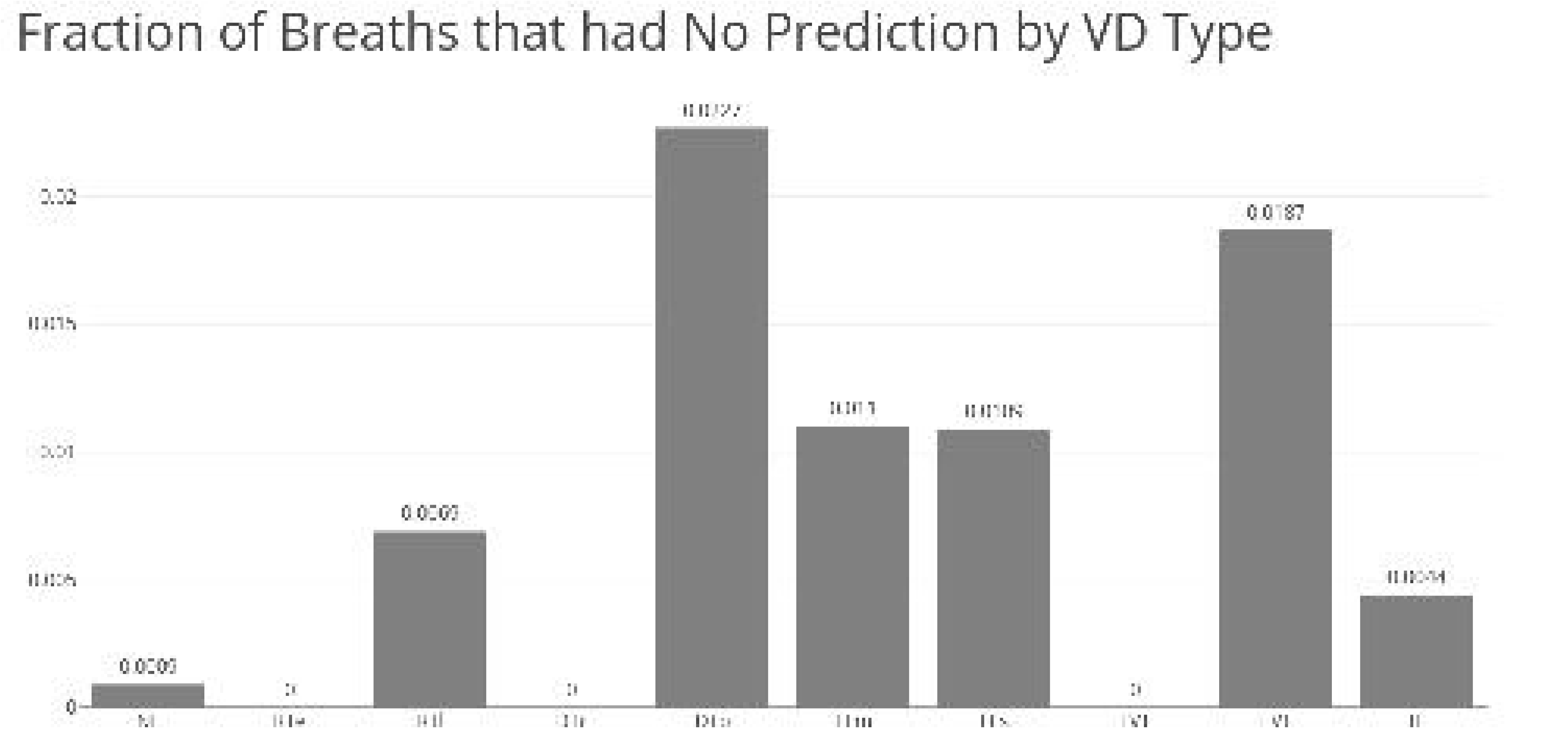
Percentage of Breaths by VD Type Left Unlabeled by XGBoost. **NL**: normal, **DT**: double-triggered, **RT**: reverse-triggered, **FL**: flow-limited; **RTe**: early reverse-triggered, **RTl**: late reverse-triggered, **DTr**: double-triggered reverse-triggered, **DTp**: patient-triggered double-triggered, **FLm**: mild flow-limited, **FLs**: severe flow-limited, **LVT**: late ventilator termination, **EVT**: early ventilator termination, **IE**: ineffective-triggered

## DISCUSSION

We created a novel computational pipeline to develop ML models that integrate esophageal pressure data and accurately label multiple types of VD on a breath-by-breath basis, including those that traditionally need advanced monitoring techniques to detect.

This paper adds to the current literature in several important ways. We devised a computational pipeline for labeling millions of breaths using ML. Identification of the VD breaths at scale is an important advancement. Refining these categories, linking these categories to physiologic changes, and using these categories to predict outcomes are all clear next steps, none of which are feasible without first being able to label breaths automatically. This algorithm will facilitate building and testing VD-specific clinical decision support tools across the 12-hospital University of Colorado Health system, leveraging our existing informatics expertise and prior experience.^39^

Developing a computational pipeline to accurately label multiple types of VD breaths in an automated fashion extends our previous work and the work of many others in three fundamentally different ways.^9,15,20–22,40–43^ First, much of the prior published work has focused on identifying only one or two types of VD in a study. We accurately predicted seven types of dyssynchrony with a single data pipeline. We could not accurately identify the RTl, EVT, and LVT breaths because of their rarity and small sample size. Second, we demonstrated the relative value of increasing model complexity to detect VD accurately. Although much work has been done previously, no study has directly compared different ML algorithms. Moreover, despite a large number of features, we did not see significant evidence of overfitting.

Considering the cost of manually labeling 10,000s of breaths and the abundance of potential features, it is important to balance model complexity with the availability of training data. Finally, and most importantly, we integrated advanced monitoring techniques (e.g., esophageal manometry) to enhance the accurate detection of multiple VD types.^18^ Pham et al. utilized p_aw_ and p_es_ data to develop a single decision to identify a breath as RT. However, this single tree was limited in identifying additional phenotypes of RT breaths, which may have a different propensity to propagate VILI. Additionally, the more complicated ensemble of decision trees utilized by XGBoost results in improved sensitivity compared to the single tree study by Pham (83.1% vs. 94.4%), again highlighting the advantage of a more complicated ML model for this classification problem.

Finally, the problem of identifying VD highlights the complexity of accurate classification in systems that generate many features but have relatively rare events. Our classification problem was characterized by complex data with significant noise, sparse outcomes, a high degree of collinearity, large datasets, and the requirement of expert knowledge to identify important key features.^29^ These problems were compounded by the cost of creating a training set through manual effort and expert knowledge. This relatively costly and small training set in the context of a highly complex classification task meant the machine learning task was complex. We observed logistic regression to be too rigid to manage the collinearity and general data complexity. The data-hungry support vector methods also faired relatively poorly. Moreover, at the extreme, while millions of mechanically ventilated breaths are generated daily, training data-hungry neural networks would not be feasible given the limitations in generating training sets. Instead, sparse data methods, such as XGBoost specifically, generally worked the best. These methods can have the risk of overfitting.^29^ To minimize this, we both manually removed features that were not informative and optimized hyperparameters to avoid overfitting. Consequently, it is not surprising that XGBoost outperformed other models.

Our study has several limitations. First, our data were collected from the University of Colorado Anschutz Medical Campus. We primarily use the Hamilton G5 ventilator and its proprietary volume target-pressure controlled mode of ventilation (adaptive pressure ventilation controlled mandatory ventilation; APVCMV). Thus, the trained models to identify VD may not be generalizable to other ventilation modes, with different features defining breath types. Second, using esophageal balloons for 48h of continuous data collection was challenging. Inaccurate or incomplete data were a significant problem that needed to be dealt with in the data cleaning process, potentially introducing systematic bias into the ML interpretation of P_es_ features because of a decrease in amplitude and magnitude of the P_es_ signal over time. Third, some types of VD were rare enough in our data set that it was difficult to obtain enough random samples for adequate ML training. Fourth, our ML models were relatively feature-rich, increasing the risk of overfitting. We attempted to adjust for this by limiting unnecessary features, utilizing regularization in the ML models, and creating an independent validation set. Finally, each VD type has substantial heterogeneity. While labeling breaths is a necessary step in understanding VD, simply labeling a breath as a type of VD will not necessarily generate a clear correlation with VILI. Further work is needed to understand this relationship.^44,45^

## CONCLUSION

We describe a novel data pipeline for the automated detection of VD, including the collection of esophageal pressure waveform data. We demonstrate that ML algorithms can be trained using esophageal pressures and airway pressure, flow, and volume to identify multiple types of VD.

This analysis highlights the need for additional work in three areas. First, streamline the data collection pathway by integrating ventilator waveforms into the EHR. Second, to simplify VD identification by extending the ML models to estimate the probability of VD type independent of esophageal manometry data. Third, to quantify the severity of VD and correlate VD with changes in pulmonary physiology to better predict which patients are harmed by VD.

## Data Availability

All data produced in the present study are available upon reasonable request to the authors and subject to approval of the University of Colorado

## ACKNOWLEDGEMENTS, COMPETING INTERESTS, FUNDING

Dr. Sottile was funded by an NIH NHLBI 5K23HL145011-05

There are no conflicts of interest reported by any authors.

## Notes

### Competing Interest Statement

The authors have declared no competing interest.

### Funding Statement

This study was funded by NIH NHLBIHL145011

### Author Declarations

The Colorado Multiple Institutional Review Board (COMIRB) gave eithical approval for this study.

