## Appendix for "The Development, Optimization, and Validation of Four Different Machine Learning Algorithms to Identify Ventilator Dyssynchrony"

**Supplemental Matierals:**

**Figure A1: Examples of Features Identified in Each Breath**

These are some of the features identified from an example RTl breath. **i1**: maximal peak inspiratory flow; **i2**: the second highest peak inspiratory flow, **e1:** the maximum peak expiratory flow; **pp**: the maximal inspiratory peak pressure; **pp_es_:** the maximal muscular contraction, most negative p_es_ deflection; **lag:** the difference between the pp and pp_es_, in seconds; dotted blue line: the width of peak i1 at 80% and 20% of the peak; gray dotted line: end of inspiration

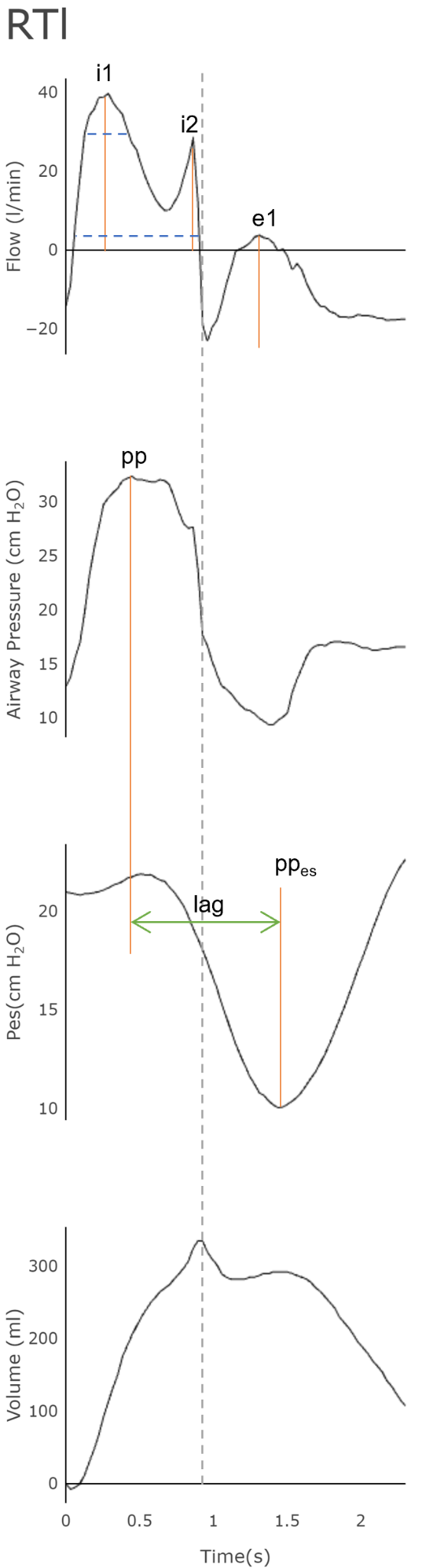

**Figure A2: Examples of Cardiac Oscillations:** Raw P_es_ is the natively recorded P_es_ tracing. Notice the frequent oscillations in the red tracing (about 3-4 a breath). After filtering, the green line results in the final P_es_ tracing for feature development. The orange box highlights a single breath. **Flow**: airway flow, **P_aw_:** airway pressure, **Vol:** volume, **P_es_:** esophageal pressure.

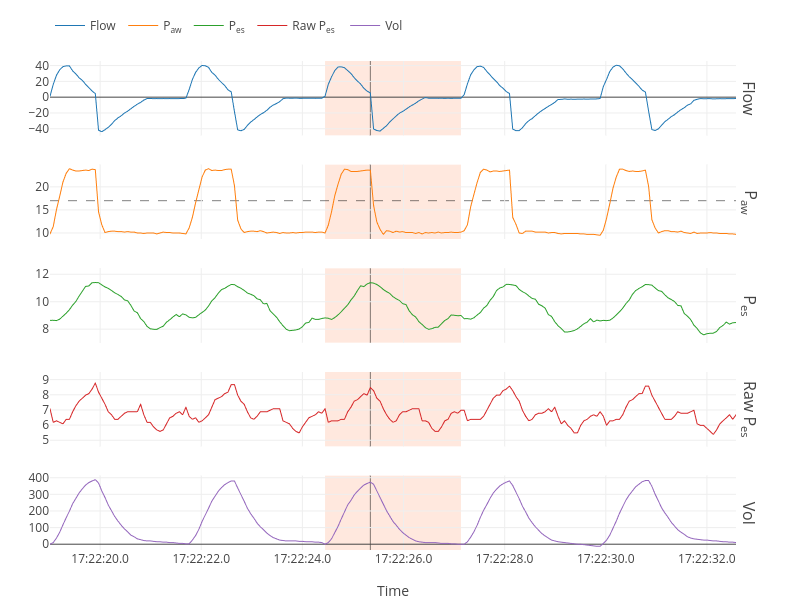

**Figures A3: Example of Violin Plots**

**A2a**: Five violin plots showing excess overlap of a given feature distribution between each type of VD. These features were excluded from ML models. *ipp_2loc*: location of the 2nd inspiratory pressure peak; *if2*: magnitude of inspiratory flow at point 2; *cove_height*: the peak of the first derivation of inspiratory p_aw_; *ipp_1ht*: the height of the 1st inspiratory peak; *epp_1ht*: the height of the first expiratory peak pressure

**A2b**: Five violin plots without excess overlap of a given feature distribution between each type of VD. These features were included in ML models. *Insp_time*: inspiratory time; *if6*: magnitude of the inspiratory flow at point 6; *paw_deviation*: the difference in measure area of inspiratory p_aw_ compared to a square weave with the same peak; *lag*: the difference between the inspiratory p_aw_ peak location and p_es_ peak location; *delta_insp_paux*: the difference between the location of peak p_es_ and end of inspiration.

**A2a**

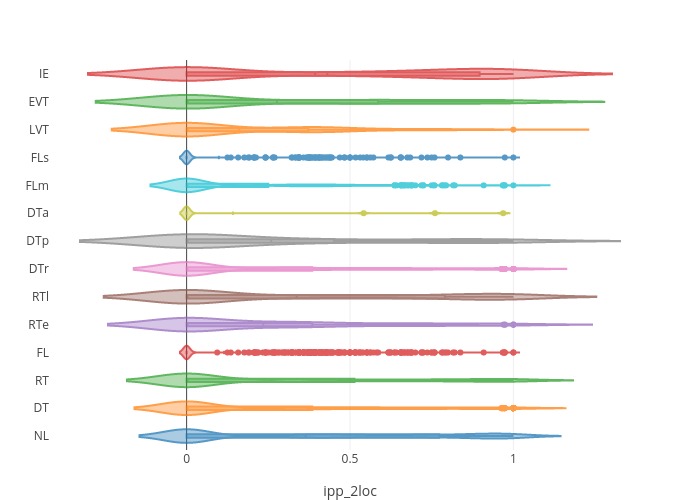

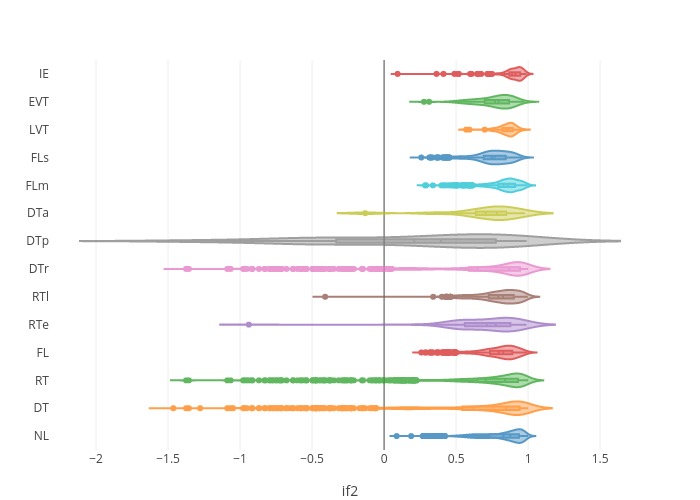

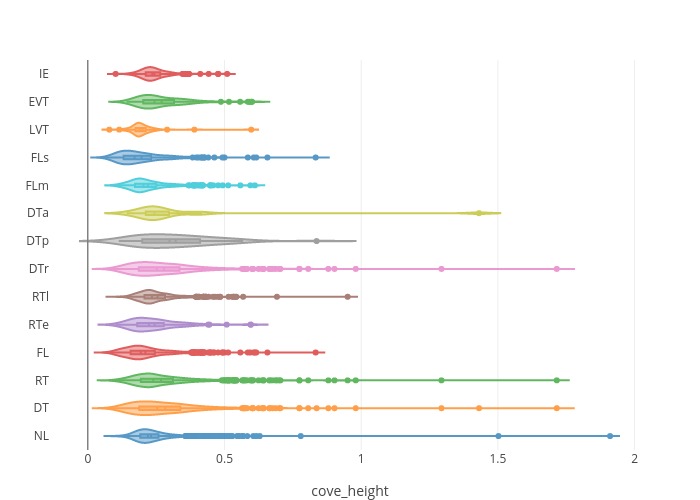

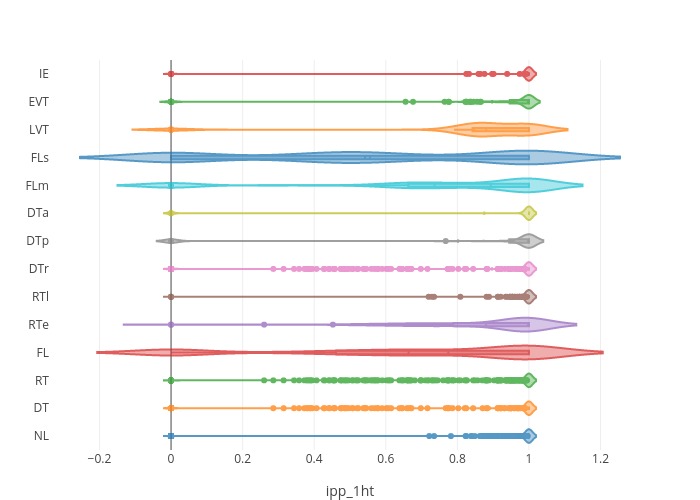

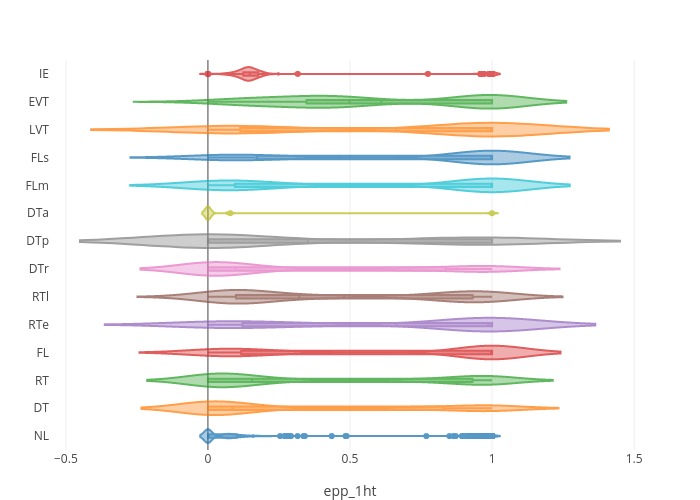

**A2b**

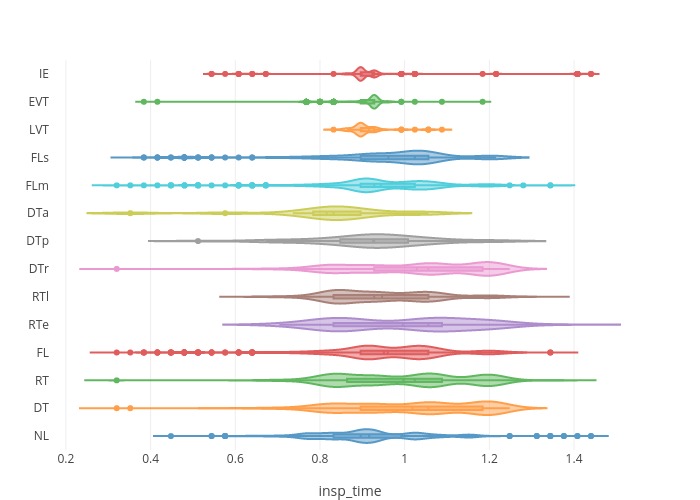

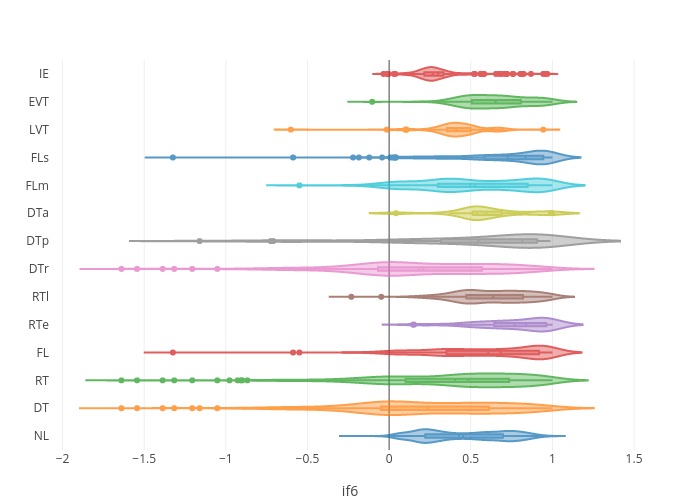

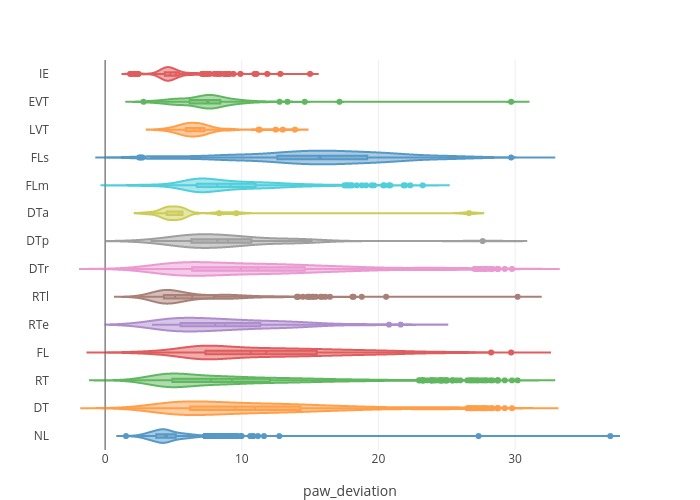

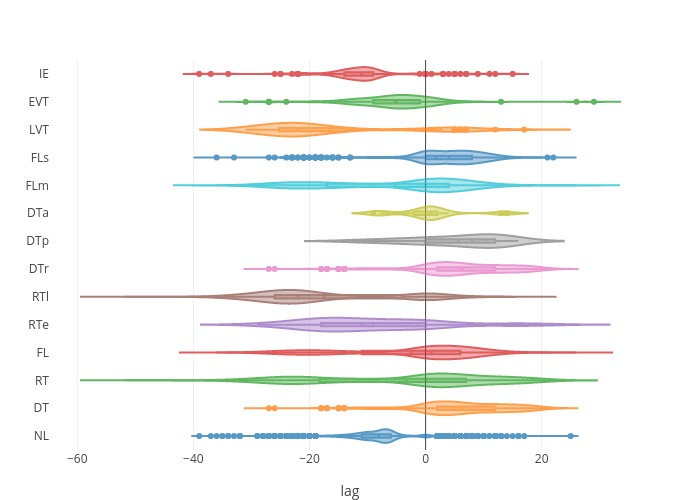

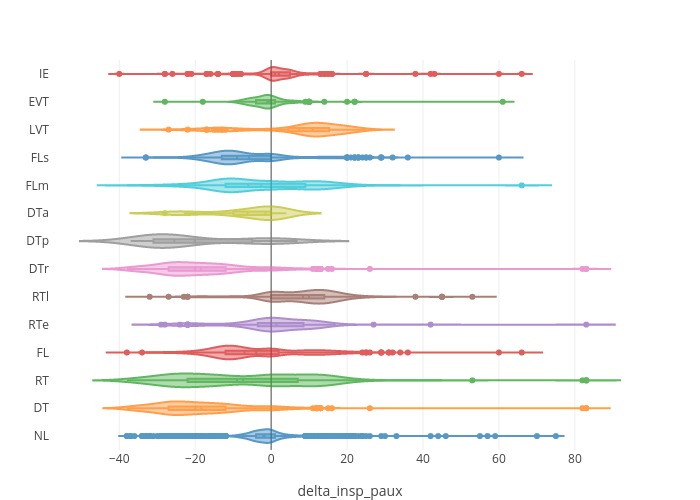

**Table A1: Test Characteristics of ML Algorithms with Esophageal Pressures:** mean % ± SD;

**Spec**: Specificity, **NL**: normal, **DT**: double-triggered, **RT**: reverse-triggered, **FL**: flow-limited; **RTe**: early reverse-triggered, **RTl**: late reverse-triggered, **DTr**: double-triggered reverse-triggered, **DTp**: patient-triggered double-triggered, **FLm**: mild flow-limited, **FLs**: severe flow-limited, **LVT**: late ventilator termination, **EVT**: early ventilator termination, **IE**: ineffective-triggered

|  | **Lasso Logistic Regression** | | | |  | **Support Vector Classification** | | | |
| --- | --- | --- | --- | --- | --- | --- | --- | --- | --- |
|  | Accuracy | F1-score | False  Negative | False  Positive |  | Accuracy | F1-score | False Negative | False  Positive |
| NL | 92.0±1.3 | 94.5±0.9 | 3.4±0.6 | 19.2±5.8 |  | 95.8±0.5 | 97.1±0.4 | 3.3±0.4 | 6.3±1.3 |
| DT | 96.3±0.6 | 73.8±5.6 | 37.1±7.7 | 0.6±0.2 |  | 98.2±0.4 | 89.1±2.2 | 14.2±3.7 | 0.6±0.3 |
| RT | 93.0±2.8 | 65.7±11.7 | 45.5±14.9 | 1.3±0.4 |  | 97.3±0.4 | 89.2±1.6 | 13.7±2.4 | 1.1±0.3 |
| FL | 94.5±0.8 | 63.9±8.5 | 45.9±10.6 | 1.4±0.4 |  | 96.8±0.3 | 81.5±2.0 | 23.1±3.0 | 1.2±0.3 |
| RTe | 99.1±0.1 | 0.0±0.0 | 1.0±0.0 | 0.0±0.0 |  | 99.1±0.1 | 18.9±9.8 | 88.4±6.4 | 0.1±0.1 |
| RTl | 98.1±1.0 | 69.4±21.9 | 37.9±25.2 | 0.4±0.2 |  | 98.6±0.2 | 82.6±3.0 | 23.1±4.2 | 0.4±.02 |
| DTr | 96.1±1.0 | 69.5±8.3 | 42.1±10.5 | 0.6±0.2 |  | 98.2±0.4 | 87.8±2.5 | 15.9±3.8 | 0.6±0.3 |
| DTp | 99.5±0.1 | 3.4±11.7 | 97.7±7.9 | 0.0±0.0 |  | 99.5±0.1 | 31.1±16.8 | 77.7±13.7 | 0.1±0.1 |
| FLm | 94.4±0.4 | 11.2±16.1 | 92.4±11.8 | 0.3±0.4 |  | 96.5±0.3 | 64.8±0.3 | 44.3±4.5 | 1.0±.2 |
| FLs | 96.7±0.4 | 21.1±23.0 | 83.9±18.8 | 0.3±0.3 |  | 98.4±0.3 | 75.2±6.0 | 31.4±7.8 | 0.5±0.2 |
| LVT | 99.3±0.1 | 0.0±0.0 | 1.0±0.0 | 0.0±0.1 |  | 99.3±.1 | 3.4±6.9 | 98.0±4.1 | 0.1±.1 |
| EVT | 98.2±0.1 | 2.9±10.1 | 98.2±7.0 | 0.1±0.1 |  | 98.8±0.3 | 60.9±10.9 | 49.0±13.4 | 0.3±0.2 |
| IE | 97.6±0.3 | 10.7±19.2 | 92.4±14.5 | 0.1±0.1 |  | 99.2±.2 | 82.9±5.1 | 25.6±7.2 | 0.1±0.1 |

|  | **Random Forest** | | | |  | **XGBoost** | | | |
| --- | --- | --- | --- | --- | --- | --- | --- | --- | --- |
|  | Accuracy | F1-score | False  Negative | False  Positive |  | Accuracy | F1-score | False Negative | False  Positive |
| NL | 96.1±0.6 | 97.2±.4 | 2.8±0.5 | 6.4±1.6 |  | 96.7±0.5 | 97.4±0.4 | 2.9±0.5 | 4.1±0.9 |
| DT | 98.70.4 | 91.6±2.3 | 12.5±3.7 | 0.3±0.1 |  | 99.2±0.22 | 94.5±1.3 | 7.4±2.3 | 0.3±0.1 |
| RT | 97.9±0.5 | 91.9±2.0 | 11.2±3.3 | 0.7±0.2 |  | 98.5±0.3 | 93.9±1.2 | 6.5±1.4 | 0.8±0.3 |
| FL | 97.8±0.4 | 89.2±1.4 | 11.3±1.8 | 1.1±0.3 |  | 98.2±0.44 | 90.1±2.0 | 10.9±2.5 | 0.9±0.3 |
| RTe | 99.0±0.2 | 14.0±11.2 | 91.8±7.2 | 0.1±0.1 |  | 99.3±0.2 | 58.6±12.5 | 50.6±13.0 | 0.2±0.1 |
| RTl | 98.9±0.2 | 88.3±2.2 | 14.2±3.8 | 0.4±.2 |  | 99.3±0.3 | 91.9±2.0 | 8.6±2.8 | 0.3±0.2 |
| DTr | 99.5±0.2 | 96.6±1.0 | 3.4±1.4 | 0.3±0.2 |  | 99.8±0.1 | 98.6±0.7 | 1.4±1.1 | 0.1±0.1 |
| DTp | 99.6±0.1 | 33.5±22.0 | 77.3±17.6 | 0.1±0.1 |  | 99.8±0.1 | 75.3±12.4 | 36.3±14.6 | 0.1±0.1 |
| FLm | 98.1±0.4 | 83.9±3.3 | 18.9±5.1 | 0.8±0.3 |  | 99.0±0.1 | 90.5±2.3 | 10.1±3.2 | 0.5±0.1 |
| FLs | 99.0±0.3 | 87.6±3.2 | 15.1±4.1 | 0.4±.2 |  | 99.1±0.3 | 88.2±3.2 | 12.3±4.9 | 0.4±0.2 |
| LVT | 99.3±.1 | 3.3±8.1 | 98.1±4.6 | 0.0±0.1 |  | 99.6±.0.1 | 54.5±15.0 | 58.2±13.7 | 0.1±0.1 |
| EVT | 99.4±.2 | 67.4±8.4 | 45.1±9.6 | 0.1±.1 |  | 99.2±0.2 | 79.7±4.8 | 29.6±6.9 | 0.1±0.1 |
| IE | 99.2±0.2 | 808±5.7 | 30.0±8.1 | 0.1±0.1 |  | 99.4±0.2 | 84.9±5.8 | 21.9±9.2 | 0.1±0.1 |
